# Operational merit of Controlled Attenuation Parameter and Liver Stiffness Measurement across grades of Metabolic Dysfunctional Associated Steatotic Liver Disease: A cross-sectional study

**DOI:** 10.1101/2025.09.28.25336846

**Authors:** S S Sujina, Poornima Ajay Manjrekar, Rajath Rao, B V Tantry, Santosh Rai, Deenadhayalan Ashok, Anupama Hegde

## Abstract

**Aim and objectives:** Metabolic dysfunction-associated steatotic liver disease (MASLD) contributes significantly to global NCD mortality. Often asymptomatic until advanced; early diagnosis prevents worsening. Given the limitations of liver biopsy, non-invasive tests (NITs) like Controlled Attenuation Parameter(CAP) and Liver Stiffness Measurement(LSM) offer promising alternatives. The present study aimed to correlate CAP with LSM and assess CAP and LSM accuracy for stratifying early steatosis and fibrosis using vibration-controlled transient elastography (VCTE).

**Methods:** The SmartExam^®^ equipment was used in patients at risk for or diagnosed with MASLD. This cross-sectional, prospective, tertiary care hospital-based study included 128 adult patients undergoing ultrasonography and/ or fibro-scan.

**Results:** Participants with steatosis grades 0-3 showed a significant increase in CAP and LSM with disease grades. A strong positive correlation existed between CAP and LSM (r=0.785, p<0.001), highlighting the interdependence of steatosis and liver stiffness. CAP demonstrated excellent diagnostic accuracy for hepatic steatosis (AUROC 1.000, p<0.001), showing substantial increases with steatosis progression. The sensitivity and specificity among individuals with high BMI/WHR were found to be 90.63% and 100% respectively, with a diagnostic accuracy of 97.66%. LSM showed marked elevation for advanced fibrosis, and the performance characteristics were found to be 81.25% sensitive, 82.29% specific, with a diagnostic accuracy of 82.03% in the higher BMI/WHR groups.

**Conclusion:** Our study confirms CAP’s excellent performance in diagnosing and stratifying hepatic steatosis and LSM’s potential role in assessing fibrosis, including early stages of MASLD. The strong CAP-LSM correlation promises synergistic utility across all stages of steatosis and liver stiffness.

## 1. Introduction

The global health landscape is increasingly dominated by Non-Communicable Diseases (NCDs), a diverse group of conditions that include type 2 diabetes mellitus (T2DM), cardiovascular diseases, cancers, hypertension, hyperlipidemia, obesity, and chronic respiratory diseases, collectively imposing an immense burden on public health systems and economies worldwide. The World Health Organization (WHO) reports that NCDs are the leading causes of mortality, accounting for over 74% of annual deaths worldwide and 63% in India(1,2). The conditions frequently co-occur, sharing common pathophysiological underpinnings and significantly elevating the risk of Metabolic dysfunction-associated steatotic Liver Disease (MASLD), which has arisen as a critical public health concern, representing the most prevalent chronic liver disease (CLD) globally and the hepatic manifestation of metabolic syndrome(3). The re-designation, from Non-alcoholic Fatty Liver Disease (NAFLD) to MASLD formalized by an international consensus in 2020 and further refined in June 2023, reflects a critical shift in understanding its pathogenesis, emphasizing its strong association with metabolic dysfunction(4). MASLD is a multisystem disease(5), often seen as a direct cause contributing to the emergence of comorbidities like T2DM, cardiovascular disease, chronic kidney diseases (CKD) and cancers(6,7). Despite being a major cause of morbidity and mortality, affecting more than 1.5 billion individuals worldwide, chronic liver disease is an under-recognized NCD that is notably absent from global NCD initiatives and frameworks.

The insidious nature of MASLD often results in delayed diagnosis or remains asymptomatic until advanced stages are reached. Therefore, early diagnosis and risk stratification help with its reversal or prevent it from further progression. the National Program for Prevention and Control of NCDs (NPNCD) by 2021 with NAFLD integration. NP-NCD had included different strategies to address this at each level of the healthcare delivery system. At the bottom level, i.e, at District hospitals(DH), fibroscans were imparted for screening(8). Although liver biopsy continues to be the definitive method for diagnosing MASLD and evaluating fibrosis severity, it has its own limitations, such as invasiveness, high cost, inconsistent histological grading, and insufficient sampling(9). Consequently, non-invasive tests (NITs) have emerged as alternatives, which include blood-based biomarkers, calculated indices and imaging techniques (10,11). However, currently, the ultrasound-based controlled attenuation parameter (CAP) and liver stiffness measurement (LSM) are widely used to assess hepatic steatosis and fibrosis in individuals with MASLD, owing to their cost-effectiveness and intuitiveness(10). The accuracy of CAP and LSM for mild steatosis (S ≥ S1), advanced liver fibrosis (F ≥ F3), and cirrhosis has been well-studied(12). The diagnostic performance of these modalities in moderate to severe steatosis and the change to fibrosis or early fibrosis in MASLD remains an area of active investigation. Understanding the intricate relationship between steatosis and fibrosis is paramount, as steatosis is often considered the “first hit” in the progression towards (Metabolic Dysfunction-Associated Steatohepatitis) MASH and subsequent fibrosis. Therefore, in keeping with the United Nations Sustainable Development Goals (SDG) goal no 3.4.1, and 12, Good Health, and wellbeing for all at all ages, to reduce premature NCD deaths through prevention and treatment and healthy food choices, reduced processed sugar/fats(13), this study aimed to check whether CAP correlates with the degree of fibrosis in terms of LSM. Also, to assess the accuracy of CAP and LSM for the stratification of early steatosis and fibrosis via vibration-controlled transient elastography (VCTE) with advanced SmartExam^®^ and automatic probe selection in a cohort of patients at risk for or diagnosed with MASLD.

## 2. Materials and methods

The study was a cross-sectional, prospective, hospital-based study conducted over a period from June 2024 to August 2025. Individuals visiting our tertiary care hospital with a requisition for abdominal ultrasonography at the department. of radiodiagnosis and fibro-scan at the dept. of Gastroenterology were included in the study upon fulfilling the inclusion criteria. Adult patients aged 18 to 65 years, of either gender, and those showing steatosis 0 to steatosis 3 on VCTE; FibroScan^®^, Echosens with advanced SmartExam and automatic probe selection, were grouped based on the Indian National Association for Study of the Liver (INASL) guidelines(14). Chronic alcoholics, as defined by the National Institute on Alcohol Abuse and Alcoholism (NIAAA) criteria(15,16)i.e.> 21 standard drinks/ week, pregnant and lactating women, individuals with ascites, T2DM patients or other secondary forms of diabetes, T2DM patients on insulin therapy, patients on lipid-lowering drugs and known cases of autoimmune hepatitis were excluded from participation(***Fig. 1***). The calculated total sample size was 128. The sampling technique used for the study was a purposive sampling technique. Ethical approval was received from the institutional ethics committee (IECKMCMLR06/2024/346). Written informed consent was obtained from all participants prior to their inclusion in the study.

**Figure 1:**
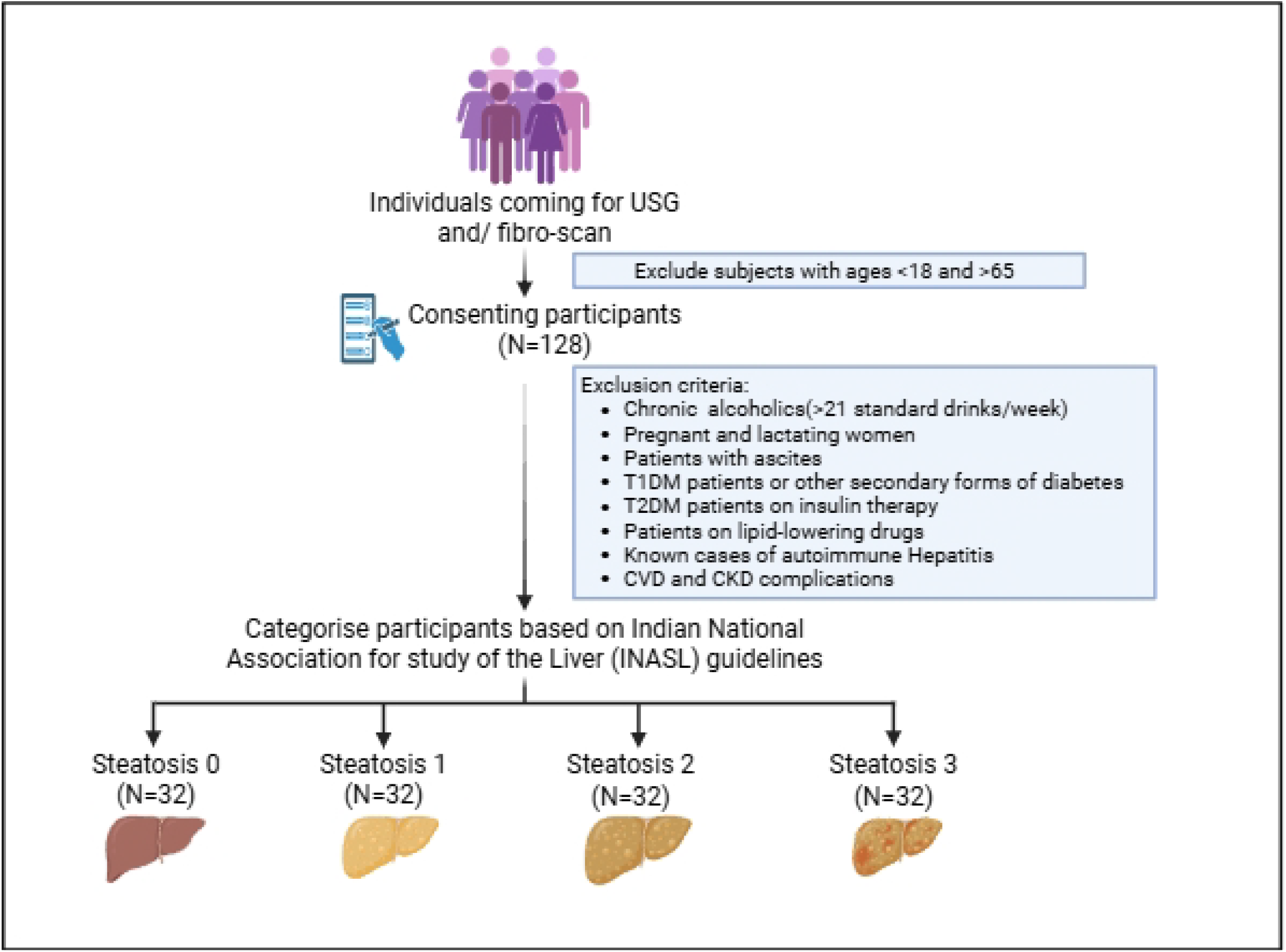
Schematic representation of study design. Created with https://BioRender.com

### 2.1. Variables and data measurements

All the variables and data used in the study were recorded by trained technicians and researchers. Steatosis grading was performed by clinicians.

#### 2.1.1. Comorbidities and lifestyle characteristics

Clinical history and lifestyle characteristics of the study participants were recorded. Physical activity was documented as per the consensus physical activity guidelines for Asian Indians(17).

#### 2.1.2. Anthropometric parameters

Height, weight, waist circumference, and hip measurements were performed via a wall-mounted height scale, digital balance scale (OMRON), and non-stretchable measuring tape, respectively, under fasting conditions according to Asian Indian population guidelines(18). Body mass index (BMI) and Waist-hip ratio(WHR) were calculated using the calculator.

#### 2.1.3. Measurement of the CAP and LSM

Steatosis and liver stiffness were measured using VCTE; FibroScan^®^, Echosens, with advanced SmartExam and automated probe selection. Following 3 hours of fasting, the examination was performed on the subjects in the supine position, with the right arm placed below the head, and the legs extended; the right leg was placed on top of the left. The selected M or XL probe (automatic probe-selection) was placed perpendicularly in the right mid-axillary line (9th–11th intercostal space) to target the right liver lobe at a depth of 25–35mm, and a minimum of 10 valid measurements were taken, with a success rate of ≥60% and an IQR/median of ≤10%. The subjects were grouped by grade as steatosis(S)/fibrosis(F) as S0/F0, S1/F1, S2/F2, and S3/F3 based on INASL guidelines(14) for CAP. The cut-off as defined by the manufacturers was S0= <248 dB/m, S1= 248 to <260 dB/m, S2= 260 to <280 dB/m, S3= ≥280 dB/m. LSM was assessed as F0-F1= 2 to 8.5KPa, and >8.5 KPa indicated fibrosis. To assess the CAP and LSM cut-offs for the increased truncal obese population, the participants were further categorized into normal (BMI ≤22.9kg/m^2^) and obese/overweight (BMI >22.9 kg/m^2^), and WHR-normal (male ≤0.9cm, female ≤0.80cm) and above-normal (male >0.9cm, female >0.80 cm) subgroups(19,20).

### 2.2. Statistical analysis

The data gathered were entered into Microsoft Excel and SPSS.29.0 (IBM Corp., Armonk, USA) was used for analysis. Descriptive statistics were applied to summarize the data. The continuous variables CAP and LSM, are reported as the Mean(SD). The categorical variables, grading (S0, S1, S2, S3 and F0, F1, F2 and F3), BMI (normal or obese/overweight) and WHR (normal/above normal) are expressed as frequencies and percentages. The associations between BMI, WHR and CAP, LSM, according to various gradings, were determined using One-way ANOVA, and a Games-Howell Post-Hoc analysis. Pearson’s correlation analysis was performed to find out the correlation between CAP and LSM. Correlation coefficient ‘r’ value is reported and a scattered plot with line of regression is plotted with the regression equation.

A receiver operating characteristic (ROC) curve was plotted for CAP and LSM with respect to various grades of steatosis and fibrosis. The area under the curve (AUC) with 95% CI and P-value is reported. The ROC curve was also plotted for the BMI sub-groups and WHR sub-groups. Specific cut-off for CAP and LSM against each grade of steatosis (S0 vs S123, S01 vs S23 and S12 vs S3) and fibrosis (F0 vs F123, F01 vs F23 and F012 vs F3) is reported with the help of Youden’s index. The respective sensitivity, specificity, positive and negative predictive values (PPV & NPV), positive and negative likelihood ratios and diagnosis accuracy (DA) were analyzed for BMI and WHR using OpenEpi software, version 3.01(21).

A multinomial logistic regression analysis was performed with grading as the dependent variable and the LSM and CAP as independent variables to predict the rates of change in the CAP and LSM with respect to various grades of steatosis and fibrosis. A crude Odds ratio with 95% CI was reported for each grading. All statistical significance was attributed to a p-value< 0.05.

## 3. Results

### 3.1) Baseline characteristics of the study participants

Among the total 128 participants, 80% were males. The mean age of the participants was 42.98(9.77) years. A small percentage of the participants had a history of diabetes (6.3%) and dyslipidemia (5.5%). Most participants were overweight or obese (88.2%) with a significant proportion (82.8%) having an abnormal waist-hip ratio (WHR). Physical activity levels were predominantly low to medium (96.9%) (**Table 1)**. Obesity and WHR is significantly (p-value<0.001) increasing in higher grades ***(S1 table)***.

**Table 1:**
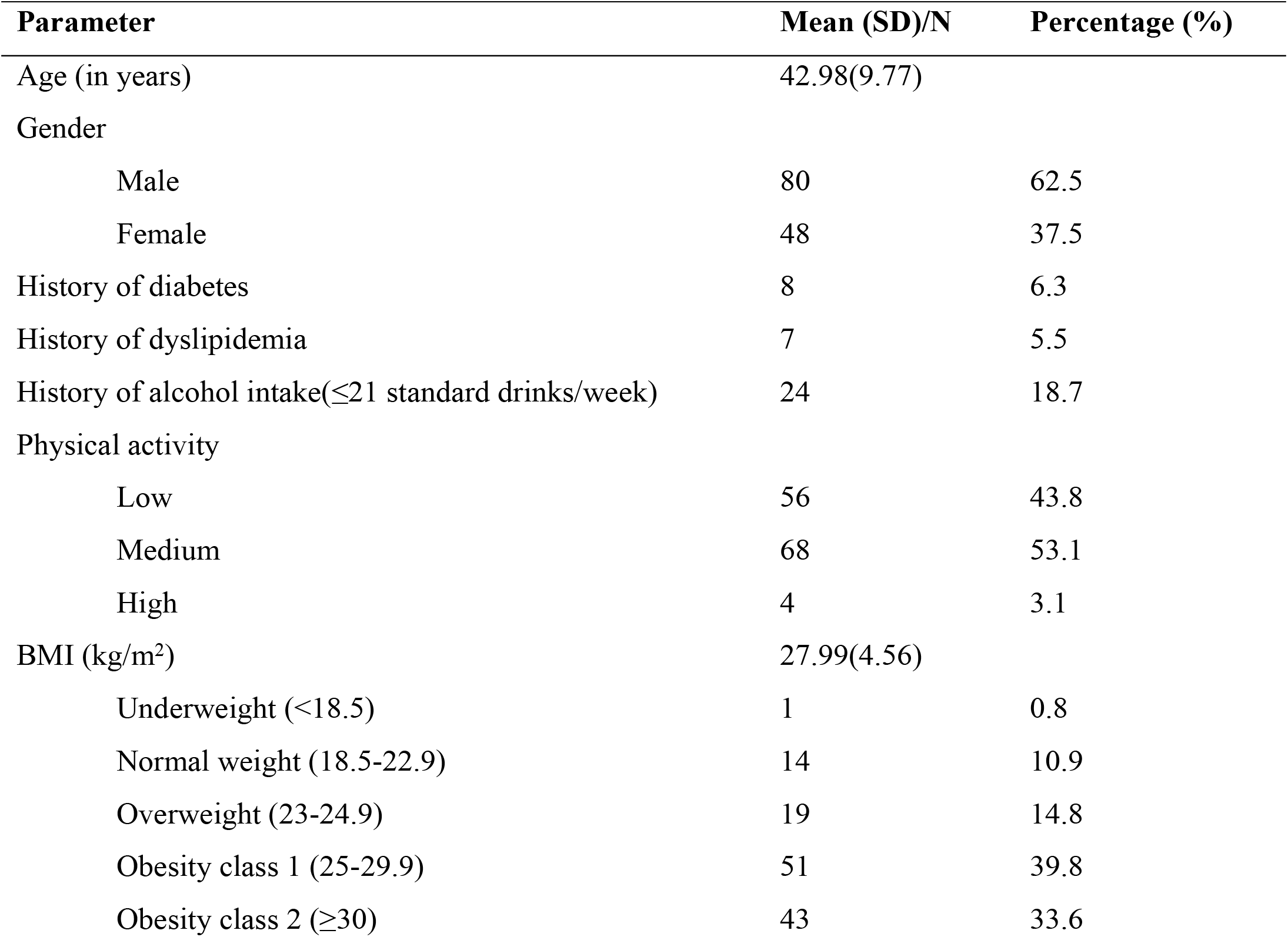
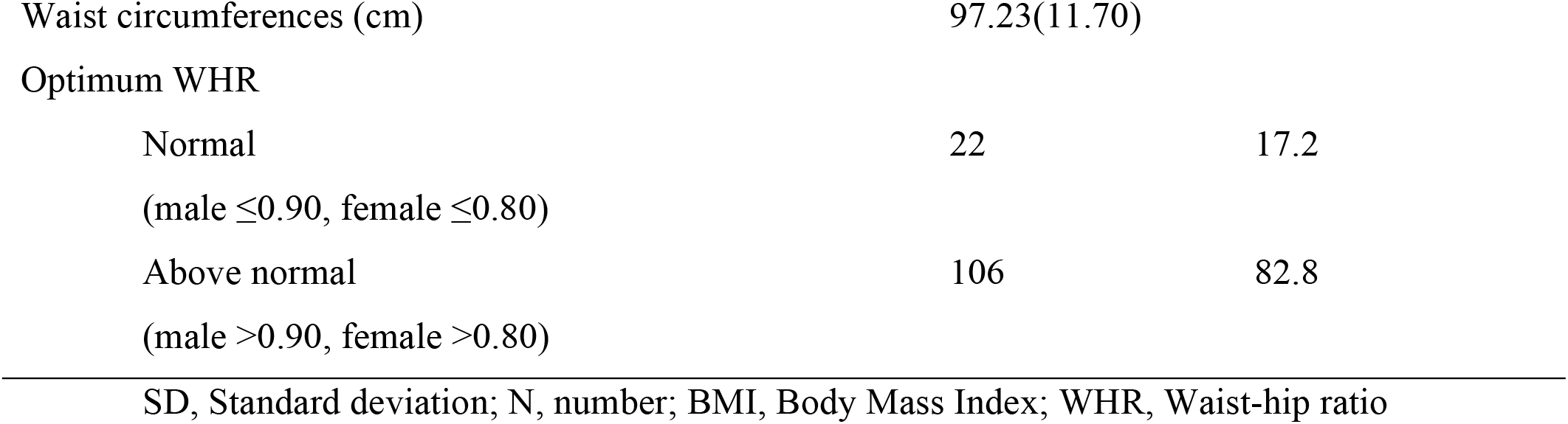
Baseline Characteristics of Study Participants (N=128)

| Parameter | Mean (SD)/N | Percentage (%) |
| --- | --- | --- |
| Age (in years) | 42.98(9.77) |  |
| Gender |  |  |
| Male | 80 | 62.5 |
| Female | 48 | 37.5 |
| History of diabetes | 8 | 6.3 |
| History of dyslipidemia | 7 | 5.5 |
| History of alcohol intake($\leq 21$ standard drinks/week) | 24 | 18.7 |
| Physical activity |  |  |
| Low | 56 | 43.8 |
| Medium | 68 | 53.1 |
| High | 4 | 3.1 |
| BMI (kg/m<sup>2</sup>) | 27.99(4.56) |  |
| Underweight (<18.5) | 1 | 0.8 |
| Normal weight (18.5-22.9) | 14 | 10.9 |
| Overweight (23-24.9) | 19 | 14.8 |
| Obesity class 1 (25-29.9) | 51 | 39.8 |
| Obesity class 2 ($\geq 30$) | 43 | 33.6 |
| Waist circumferences (cm) | 97.23(11.70) |  |
| Optimum WHR |  |  |
| Normal | 22 | 17.2 |
| (male $\leq 0.90$, female $\leq 0.80$) | | |
| Above normal | 106 | 82.8 |
| (male $> 0.90$, female $> 0.80$) | | |
| SD, Standard deviation; N, number; BMI, Body Mass Index; WHR, Waist-hip ratio |  |  |

### 3.2) Comparison of CAP and LSM across grades of MASLD

***Table 2*** delineates the comparison of CAP and LSM of the study participants in different grades of MASLD. The mean CAP and LSM for all participants revealed a consistent and statistically significant increase with advancing steatosis grades (p<0.001), reflecting heightened liver fat content and progressive fibrosis, respectively. In subgroup analysis based on BMI and WHR, individuals classified as obese/overweight (n=113) and above normal WHR (n=106) displayed slightly higher mean CAP values and LSM values across the grades. Statistical analysis revealed a strong significant difference in CAP and LSM values at any stage (p<0.001) except for normal BMI group (p=0.006). **(Table 2)**.

**Table 2:**
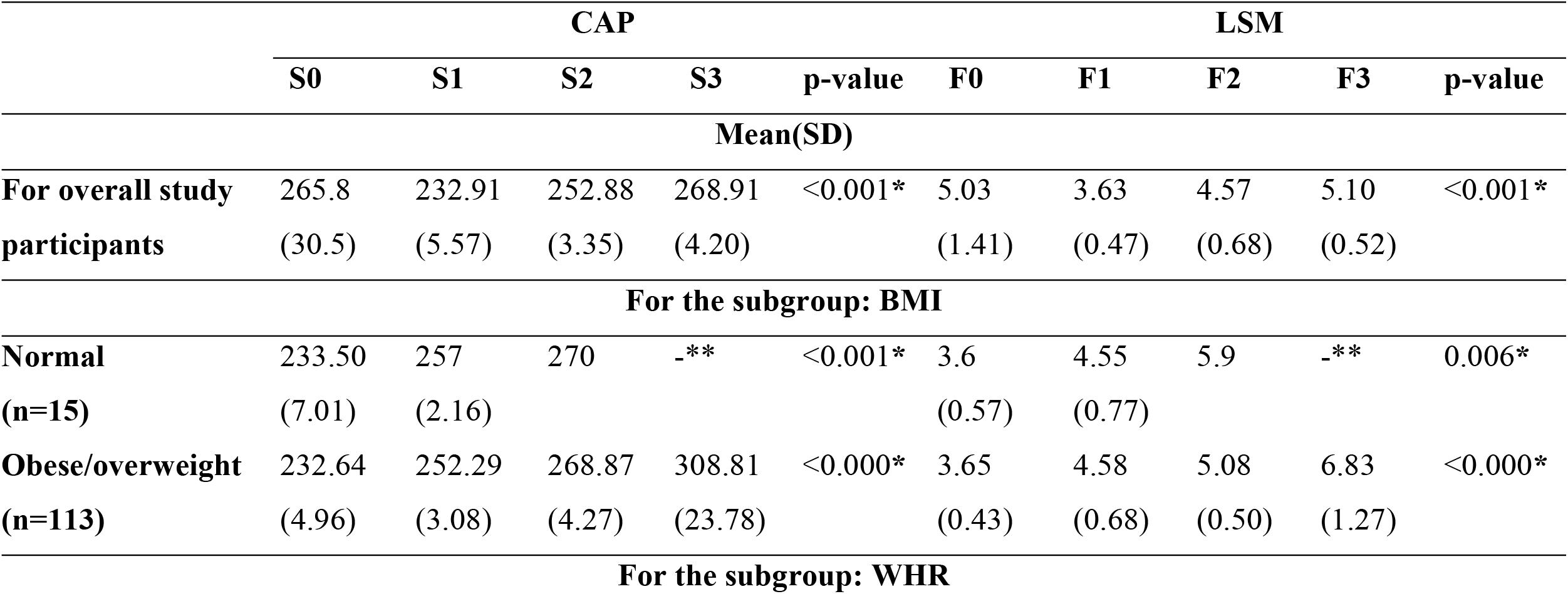
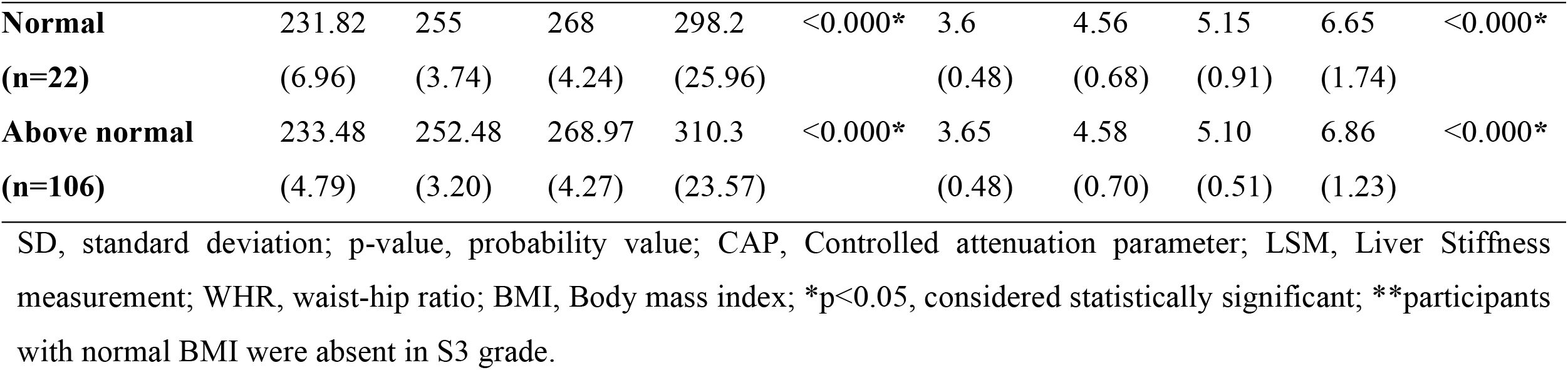
CAP and LSM of the study participants (N=128)

| Table 2: CAP and LSM of the study participants (N=128) |  |  |  |  |  |  |  |  |  |  |
| --- | --- | --- | --- | --- | --- | --- | --- | --- | --- | --- |
|  | CAP |  |  |  |  | LSM |  |  |  |  |
|  | S0 | S1 | S2 | S3 | p-value | F0 | F1 | F2 | F3 | p-value |
| Mean(SD) |  |  |  |  |  |  |  |  |  |  |
| **For overall study participants** | 265.8<br>(30.5) | 232.91<br>(5.57) | 252.88<br>(3.35) | 268.91<br>(4.20) | <0.001* | 5.03<br>(1.41) | 3.63<br>(0.47) | 4.57<br>(0.68) | 5.10<br>(0.52) | <0.001* |
| **For the subgroup: BMI** |  |  |  |  |  |  |  |  |  |  |
| **Normal (n=15)** | 233.50<br>(7.01) | 257<br>(2.16) | 270 | -** | <0.001* | 3.6<br>(0.57) | 4.55<br>(0.77) | 5.9 | -** | 0.006* |
| **Obese/overweight (n=113)** | 232.64<br>(4.96) | 252.29<br>(3.08) | 268.87<br>(4.27) | 308.81<br>(23.78) | <0.000* | 3.65<br>(0.43) | 4.58<br>(0.68) | 5.08<br>(0.50) | 6.83<br>(1.27) | <0.000* |
| **For the subgroup: WHR** |  |  |  |  |  |  |  |  |  |  |
| **Normal** | 231.82 | 255 | 268 | 298.2 | <0.000* | 3.6 | 4.56 | 5.15 | 6.65 | <0.000* |
| **(n=22)** | (6.96) | (3.74) | (4.24) | (25.96) |  | (0.48) | (0.68) | (0.91) | (1.74) |  |
| **Above normal** | 233.48 | 252.48 | 268.97 | 310.3 | <0.000* | 3.65 | 4.58 | 5.10 | 6.86 | <0.000* |
| **(n=106)** | (4.79) | (3.20) | (4.27) | (23.57) |  | (0.48) | (0.70) | (0.51) | (1.23) |  |

SD, standard deviation; p-value, probability value; CAP, Controlled attenuation parameter; LSM, Liver Stiffness measurement; WHR, waist-hip ratio; BMI, Body mass index; \*p<0.05, considered statistically significant; \*\*participants with normal BMI were absent in S3 grade.

### 3.3) Correlation between overall CAP and LSM

A Pearson’s correlation was conducted to assess the relationship between Controlled Attenuation Parameter (CAP) and Liver Stiffness Measurement (LSM). The results demonstrated a strong positive significant correlation (r=0.785, p value <0.001) was noted with an R^2^ value of 0.616, indicating that approximately 61.6% of the variance in LSM values could be explained by CAP. The regression equation obtained was: LSM = 0.051 × CAP – 10. ***(Figure 2)***

**Figure 2:**
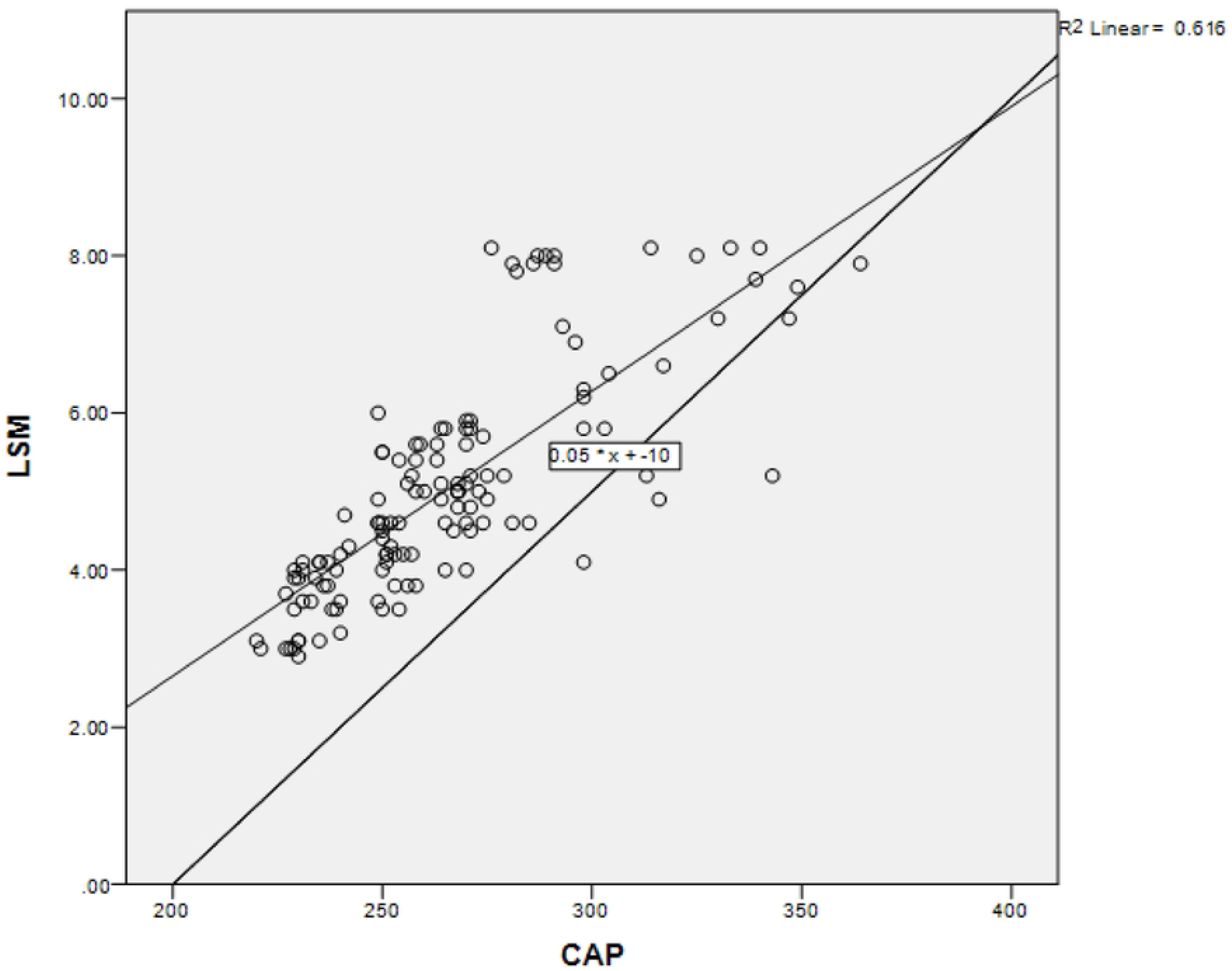
Scatter plot showing the correlation between Controlled Attenuation Parameter (CAP) and Liver Stiffness Measurement (LSM) in study participants. Each point represents an individual subject’s CAP (dB/m) and corresponding LSM (kPa) values. A linear regression line is overlaid (solid line), indicating a positive linear relationship between CAP and LSM. The regression equation derived is LSM = 0.051 × CAP – 10, with a coefficient of determination R^2^ = 0.616, suggesting that 61.6% of the variability in LSM is explained by CAP values. The diagonal reference line (dashed) represents the identity line (y = x) for visual comparison. This analysis highlights the potential interdependence between hepatic steatosis and fibrosis in the MASLD spectrum.

### 3.4) Diagnostic characteristics of CAP and LSM

***Table 3*** depicts the overview of diagnostic characteristics of CAP for the diagnosis of hepatic steatosis in the study cohort. The AUROC (95%CI) for CAP was 1.000(1.000-1.000), (p<0.001) indicating efficient predictive capability in distinguishing between S0 v/s S123, S01 v/s S23 and S012 v/s S3. The cut-off values for detecting steatosis ≥ S1, ≥ S2 and ≥ S3 were found to be 245.5 dB/m, 259.5 dB/m and 275.5 dB/m respectively ***(fig. 3 A-C)***. These cut-offs were 100% sensitive and 100% specific. Similarly, in sub-group analysis of WHR normal and WHR above normal, CAP maintained a high diagnostic performance (AUROC, 95% CI= 1.000(1.000-1.000)) with 100% sensitivity and specificity with similar values ***(fig. 3 D,E and I-K)***.

**Table 3:** Diagnostic characteristics of CAP for the diagnosis of hepatic steatosis among MASLD cohort (N=128).

|  | AUROC | Cut-off | Sensitivity | Specificity | Positive predictive value | Negative predictive value | Diagnostic accuracy | p-value |
| --- | --- | --- | --- | --- | --- | --- | --- | --- |
|  |  |  |  |  | (95%CI) |  |  |  |
| **For overall study participants, n= 128** |  |  |  |  |  |  |  |  |
| S0 vs S123 | 1.00<br>(1.00-1.00) | 245.5 | 100<br>(96.15- 100) | 100<br>(89.28- 100) | 100%<br>(95.15-100) | 100%<br>(89.28- 100) | 100%<br>(97.09-100) | <0.001* |
| S01 vs S23 | 1.00<br>(1.00-1.00) | 259.5 | 100<br>(93.12-100) | 100<br>(94.34-100) | 100%<br>(94.34-100) | 100%<br>(94.34-100) | 100%<br>(97.09-100) | <0.001* |
| S012 vs S3 | 1.00<br>(1.00-1.00) | 275.5 | 100<br>(89.28-100) | 98.96<br>(94.33-99.82) | 96.97%<br>(84.68- 99.46) | 100%<br>(96.11-100) | 99.22% (95.71-99.86) | <0.001* |
| **For the subgroup BMI** |  |  |  |  |  |  |  |  |
| **BMI normal (BMI ≤22.9kg/m<sup>2</sup>), n= 15** |  |  |  |  |  |  |  |  |
| S0 vs S123 | 1.00<br>(1.00-1.00) | 241.5 | 100<br>(96.15-100) | 96.88%<br>(84.26- 99.45) | 98.97% (94.39-99.82) | 100%<br>(88.97-100) | 99.22% (95.71-99.86) | 0.002* |
| S01 vs S23 | 1.00<br>(1.00-1.00) | 258.5 | 100<br>(94.34-100) | 96.88%<br>(89.3- 99.14) | 96.97%<br>(89.61- 99.17) | 100%<br>(94.17-100) | 98.44% (94.48-99.57) | 0.105 |
| S012 vs S3** | - | - | - | - | - | - | - | - |
| **Obese/ overweight (BMI >22.9kg/m<sup>2</sup>), n= 113** |  |  |  |  |  |  |  |  |
| S0 vs S123 | 1.0<br>(1.0-1.0) | 249.5 | 94.79%<br>(88.38-97.76) | 100%<br>(89.28- 100) | 100%<br>(95.95-100) | 86.49%<br>(72.02-94.09) | 96.09% (91.18-98.32) | <0.001* |
| S01 vs S23 | 1.0<br>(1.0-1.0) | 264.5 | 90.63%<br>(81.02-95.63) | 100%<br>(94.34- 100) | 100%<br>(93.79- 100) | 91.43%<br>(82.53-96.01) | 95.31% (90.15- 97.83) | <0.001* |
| S012 vs S3 | 1.0<br>(0.99-1.0) | 281.5 | 90.63%<br>(75.78-96.76) | 100%<br>(96.15- 100) | 100%<br>(88.3- 100) | 96.97%<br>(91.47-98.96) | 97.66% (93.34- 99.2) | <0.001* |
| **For the subgroup WHR** |  |  |  |  |  |  |  |  |
| **WHR Normal, n= 22** |  |  |  |  |  |  |  |  |
| S0 vs S123 | 1.0<br>(1.0- 1.0) | 245 | 100%<br>(96.15- 100) | 100%<br>(89.28- 100) | 100%<br>(96.15- 100) | 100%<br>(89.28- 100) | 100%<br>(97.09- 100) | <0.001* |
| S01 vs S23 | 1.0<br>(1.0- 1.0) | 257.5 | 100%<br>(94.34- 100) | 92.19%<br>(82.98- 96.62) | 92.75%<br>(84.13- 96.87) | 100%<br>(93.89- 100) | 96.09%<br>(91.18 98.32) | <0.001* |
| S012 vs S3 | 1.0<br>(1.0- 1.0) | 273.5 | 100%<br>(89.28- 100) | 94.79%<br>(88.38- 97.76) | 86.49%<br>(72.02- 94.09) | 100%<br>(95.95- 100) | 96.09% (91.18- 98.32) | 0.002* |
| **WHR above normal, N=106** |  |  |  |  |  |  |  |  |
| S0 vs S123 | 1.0<br>(1.0- 1.0) | 249.5 | 95%<br>(88.38- 97.76) | 100%<br>(89.28-100) | 100%<br>(95.95-100) | 86.49%<br>(72.02-94.09) | 96%<br>(91.18-98.32) | <0.001* |
| S01 vs S23 | 1.0<br>(1.0- 1.0) | 263.5 | 95.31%<br>(87.1-98.39) | 100%<br>(94.34-100) | 100%<br>(94.08-100) | 95.52%<br>(87.64-98.47) | 97.66% (93.34- 99.2) | <0.001* |
| S012 vs S3 | 1.0<br>(1.0- 1.0) | 281.5 | 90.63%<br>(75.78-96.76) | 100%<br>(96.15-100) | 100%<br>(88.3, 100) | 96.97%<br>(91.47-98.96) | 97.66% (93.34- 99.2) | <0.001* |

AUROC, area under receiver operator characteristic curve; MASLD, metabolic associated steatotic liver disease; CI, confidence interval; p-value, probability value; S, steatosis, WHR, waist-hip ratio; \*p<0.05, considered statistically significant; \*\*participants with normal BMI were absent in S3 grade.

**Figure 3:**
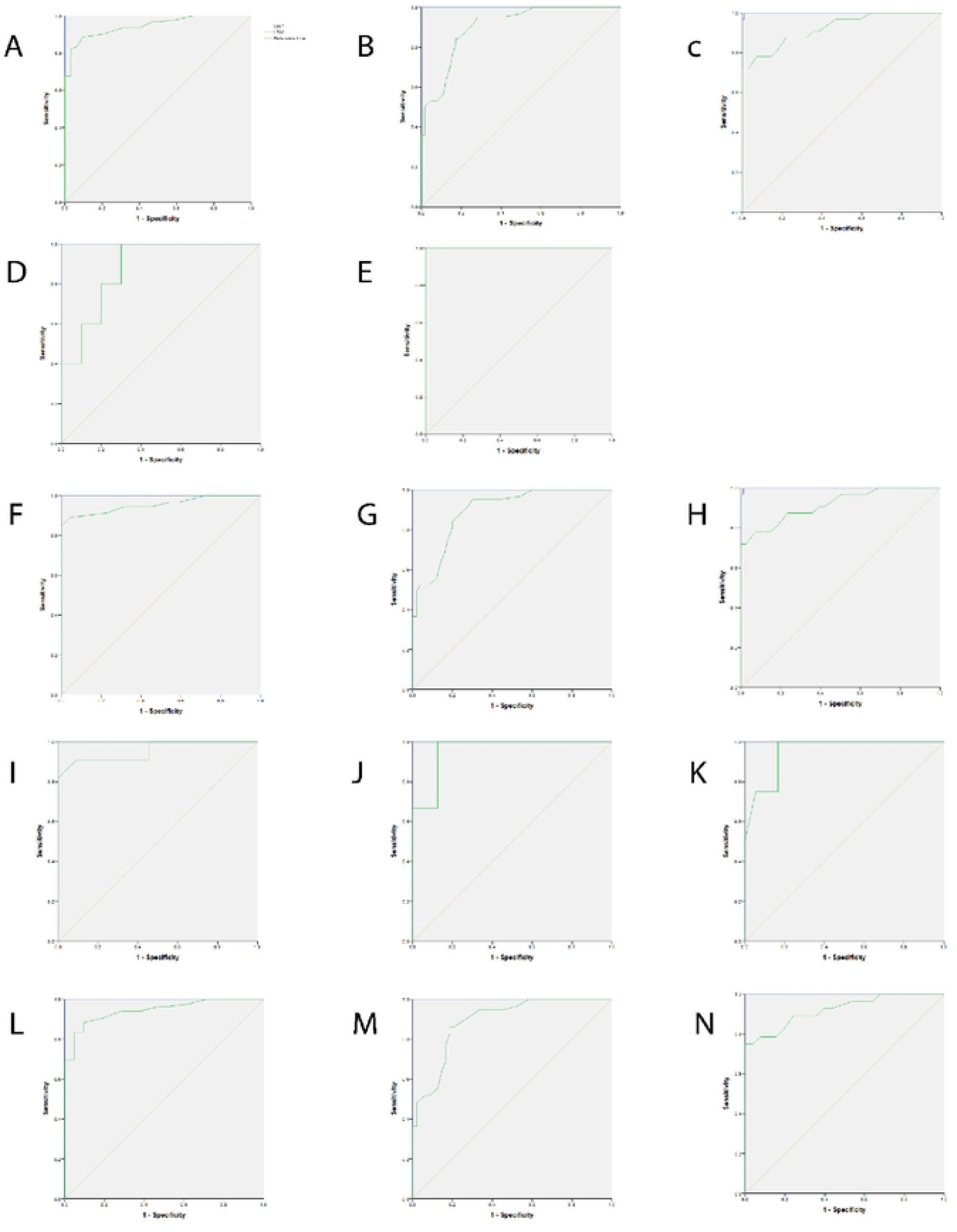
From: Imaging parameters: CAP and LSM for measuring steatosis and fibrosis in study participants. Receiver operating characteristic (ROC) curves of controlled attenuation parameter (CAP) and liver stiffness measurement (LSM), whole study participants, A S0 v/s S123, B S01 v/s S23, C S012 v/s S3. ROC curves of CAP and LSM, BMI normal, D S0 v/s S123, E S01 v/s S23. ROC curves of CAP and LSM, Obese/overweight, F S0 v/s S123, G S01 v/s S23, H S012 v/s S3, ROC curves of CAP and LSM, WHR normal, I S0 v/s S123, J S01 v/s S23, K S012 v/s S3. ROC curves of CAP and LSM, WHR above normal, L S0 v/s S123, M S01 v/s S23, N S012 v/s S3.**H** S012 v/s S3.

In individuals with obesity/ overweight and WHR-above normal, the cut-offs for detecting ≥ S1, ≥ S2, and S3 were 249.5 dB/m, 264.5 dB/m, and 281.5 dB/m (p<0.001) ***(fig 3 F-H)*** and 249.5 dB/m, 263.5 dB/m, and 281 dB/m (p<0.001)) ***(fig 3 L-N)*** respectively. **(Table 3)**

The diagnostic performance of LSM ***(Table 4)***, with an AUROC (95%CI) for detecting ≥ F1, ≥ F2, and F3 was 0.94 (0.91-0.98), 0.90 (0.85-0.95), and 0.92 (0.86-0.98), respectively. The cut-off to distinguish between ≥ F1, ≥ F2, and F3 was 3.95 kPa, 4.45 kPa and 5.05 kPa (p<0.000) ***(fig 3 A-C)*** respectively. The sensitivity, specificity, PPV, NPV, and DA of various cut-offs of CAP are shown in ***Table 4***. In the normal BMI and WHR subgroups, F0 was found to be 3.9 kPa ***(fig 3 D, E and I-K)*** with a diagnostic accuracy of 87.50% for both the subgroups. This cutoff was increased by 0.25 kPa and 0.15 kPa ***(fig 3 F-H &L-N)*** in obese/ overweight and WHR above normal subgroups for detecting ≥ F1, ≥ F2, and F3. **(Table 4)**

**Table 4:** Diagnostic characteristics of LSM for the diagnosis of liver stiffness among the MASLD cohort (N=128).

|  | AUROC | Cut-off | Sensitivity | Specificity | Positive<br>predictive<br>value | Negative<br>predictive<br>value | Diagnostic<br>accuracy | p-value |
| --- | --- | --- | --- | --- | --- | --- | --- | --- |
| (95%CI) |  |  |  |  |  |  |  |  |
| **For overall study participants, n=128** |  |  |  |  |  |  |  |  |
| F0 vs F123 | 0.94<br>(0.91-0.98) | 3.95 | 93.75%<br>(87.03- 97.1) | 68.75%<br>(51.43- 82.05) | 90%<br>(82.56- 94.48) | 78.57%<br>(60.46-89.79) | 87.50%<br>(80.66- 92.16) | <0.001<br>* |
| F01 vs F23 | 0.90<br>(0.85-0.95) | 4.45 | 95.31%<br>(87.1- 98.39) | 71.88%<br>(59.87- 81.41) | 77.22%<br>(66.83-85.07) | 93.88%<br>(83.48-97.9) | 83.59%<br>(76.22- 89.01) | <0.001<br>* |
| F012 vs F3 | 0.92<br>(0.86-0.98) | 5.05 | 87.50%<br>(71.93- 95.03) | 73.96%<br>(64.38- 81.69) | 52.83%<br>(39.66- 65.62) | 94.67%<br>(87.07-97.91) | 77.34%<br>(69.36-83.74) | <0.001<br>* |
| **For the subgroup BMI** |  |  |  |  |  |  |  |  |
| **BMI ≤22.9kg/m<sup>2</sup>, n=15** |  |  |  |  |  |  |  |  |
| F0 vs F123 | 0.88<br>(0.70-1.00) | 3.9 | 93.75%<br>(87.03- 97.1) | 68.75%<br>(51.43- 82.05) | 90% (82.56-94.48) | 78.57%<br>(60.46-89.79) | 87.50%<br>(80.66- 92.16) | 0.020* |
| F01 vs F23 | 1.00<br>(1.00-1.00) | 4.4 | 95.31% (87.1-98.39) | 71.88%<br>(59.87- 81.41) | 77.22%<br>(66.83- 85.07) | 93.88%<br>(83.48- 97.9) | 83.59%<br>(76.22-89.01) | 0.105 |
| F012 vs F3** | - | - | - | - | - | - | - |  |
| **BMI >22.9kg/m<sup>2</sup>, n=113** |  |  |  |  |  |  |  |  |
| F0 vs F123 | 0.95<br>(0.92-0.98) | 4.15 | 88.54%<br>(80.64- 93.48) | 90.63%<br>(75.78-96.76) | 96.59%<br>(90.45-98.83) | 72.50%<br>(57.16-83.89) | 89.06%<br>(82.48- 93.37) | <0.001<br>* |
| F01 vs F23 | 0.89<br>(0.83-0.95) | 4.7 | 84.38%<br>(73.57-91.29) | 82.81%<br>(71.79-90.12) | 83.08%<br>(72.18-90.28) | 84.13%<br>(73.19-91.14) | 83.59%<br>(76.22-89.01) | <0.001<br>* |
| F012 vs F3 | 0.91<br>(0.85-0.97) | 5.3 | 81.25%<br>(64.69- 91.11) | 82.29%<br>(73.46- 88.64) | 60.47%<br>(45.58- 73.63) | 92.94%<br>(85.44-96.72) | 82.03%<br>(74.48- 87.72) | <0.001<br>* |
| **For the subgroup WHR** |  |  |  |  |  |  |  |  |
| **WHR Normal, n= 22** |  |  |  |  |  |  |  |  |
| F0 vs F123 | 0.95<br>(0.86-1.00) | 3.9 | 93.75%<br>(87.03-97.1) | 68.75%<br>(51.43-82.05) | 90%<br>(82.56- 94.48) | 78.57%<br>(60.46-89.79) | 87.50%<br>(80.66-92.16) | <0.001<br>* |
| F01 vs F23 | 0.95<br>(0.88-1.0) | 4.4 | 95.31%<br>(87.1-98.39) | 71.88%<br>(59.87- 81.41) | 77.22%<br>(66.83- 85.07) | 93.88%<br>(83.48- 97.9) | 83.59%<br>(76.22- 89.01) | <0.001<br>* |
| F012 vs F3 | 0.95<br>(0.85-1.00) | 4.8 | 90.63%<br>(75.78- 96.76) | 64.58%<br>(54.62- 73.42) | 46.03%<br>(34.31- 58.21) | 95.38%<br>(87.29-98.42) | 71.09%<br>(62.72- 78.24) | 0.006* |
| **WHR above normal, N=106** |  |  |  |  |  |  |  |  |
| F0 vs F123 | 0.94<br>(0.90-0.98) | 4.05 | 90.63%<br>(83.13-94.99) | 78.13%<br>(61.24- 88.98) | 92.55%<br>(85.42-96.35) | 73.53%<br>(56.88-85.4) | 87.50%<br>(80.66-92.16) | <0.001<br>* |
| F01 vs F23 | 0.89<br>(0.83-0.95) | 4.75 | 84.38%<br>(73.57-91.29) | 82.81%<br>(71.79-90.12) | 83.08%<br>(72.18-90.28) | 84.13%<br>(73.19-91.14) | 83.59%<br>(76.22-89.01) | <0.001<br>* |
| F012 vs F3 | 0.92<br>(0.85-0.98) | 5.3 | 81.25%<br>(64.69-91.11) | 82.29%<br>(73.46-88.64) | 60.47%<br>(45.58- 73.63) | 92.94%<br>(85.44-96.72) | 82.03%<br>(74.48-87.72) | <0.001<br>* |

AUROC, area under receiver operator characteristic curve; MASLD, metabolic associated steatotic liver disease; CI, confidence interval; p-value, probability value; WHR, waist-hip ratio; F: fibrosis; \*p<0.05, considered statistically significant; \*\*participants with normal BMI were absent in S3 grade.

### 3.4) Rate of change of CAP and LSM across various grades of MASLD

Multinomial logistic regression analysis ***(table 5)*** was done to find out the rate of change of the imaging parameters with respect to various stages of MASLD. Compared to S0, the CAP values were 1.97 times (COR=1.97, 95% CI=1.97-1.99, p<0.001) more in S1, 5.05 times (COR=5.05, 95% CI=4.99-5.11, p<0.001) in S2 and 8.8 times (COR=8.8, 95% CI=8.69-8.91, p<0.001) in S3, indicating a substantial increase in the likelihood of severe steatosis. Similarly, LSM demonstrated a marked elevation in mean values from F0 to F3, with crude odds ratios indicating an even more pronounced increase in the probability of advanced fibrosis, reaching 453.2 for F3 relative to F0. **(Table 5)**

**Table 5:** Multinominal logistic regression table for predicting the rate of change of CAP and LSM across various grades (N=128).

|  |  | Mean (SD) | Crude OR<br>(95% CI) | p-value |
| --- | --- | --- | --- | --- |
| CAP, dB/m | S0 | 232.91(5.57) | Ref. |  |
|  | S1 | 252.88(3.35) | 1.97(1.97-1.99) | <0.001* |
|  | S2 | 268.91(4.20) | 5.05(4.99- 5.11) | <0.001* |
|  | S3 | 308.81(23.78) | 8.80(8.69-8.91) | <0.001* |
| LSM, kPa | F0 | 3.63(0.47) | Ref. |  |
|  | F1 | 4.57(0.68) | 26.9(5.61-126) | <0.001* |
|  | F2 | 5.10(0.52) | 89.4(15.95-502) | <0.001* |
|  | F3 | 6.83(1.27) | 453.2(69.5-2955) | <0.001* |

SD, standard deviation; OR, Odds ratio; CI, confidence intervals; p-value, probability value; CAP, Controlled Attenuation Parameter; LSM, liver stiffness measurement; Ref. reference; \*p<0.05, considered statistically significant.

## Discussion

The findings of the present study highlight a high DA of CAP for detecting hepatic steatosis in MASLD patients, demonstrating 100% sensitivity and specificity with excellent AUROC values across different steatosis grades (≥S1, ≥S2, and S3). Our results, with CAP cut-off values of 245.5 dB/m for ≥S1, 259.5 dB/m for ≥S2, and 275.5 dB/m for S3, are consistent with previous research that validates CAP as a reliable non-invasive tool for quantifying liver steatosis(22,23). The results were slightly lower than the manufacturer’s cutoffs. CAP maintained a high diagnostic performance even in subgroups with normal BMI and WHR with similar values. It is important to acknowledge that the accuracy of CAP for the detection of steatosis in patients with BMI and WHR above normal values is slightly reduced, with higher cut-off values. Shalima et. al reported higher cutoffs for BMI> 30 kg/m^2^ and presented the importance of the use of specific probes(24). However, even after the use of automated probe selection, slightly elevated cutoffs for overweight and above normal WHR subgroups indicate that CAP must be combined with some other non-invasive tests (NITs), for 100% accuracy in diagnosing steatosis in such patients for clinical diagnosis.

While CAP demonstrated superior performance for steatosis, the diagnostic performance of LSM for fibrosis was slightly lower, with AUROC values for ≥F1, ≥F2, and F3 being 0.94, 0.90, and 0.92, respectively. The corresponding cut-offs were 3.95 kPa, 4.45 kPa, and 5.05 kPa. These LSM values, while indicative of fibrosis, suggest that LSM may have slightly reduced discriminatory power compared to CAP in distinguishing between early fibrosis stages in our cohort. Fujiwara et al. confirm that LSM is a promising tool for noninvasive quantification of hepatic fibrosis(25). Similar to our study, Cao et al. have also stated LSM values may be used for the stratification of fibrotic stages(26), highlighting the possible role of LSM in distinguishing between early and established fibrosis. However, studies suggest that LSM may be less accurate in patients with severe obesity (BMI ≥ 35 kg/m^2^)(27). The AUROC of LSM for detecting ≥ F1, ≥ F2, and ≥ F3 in obese/ overweight and WHR above normal cohort were 0.95, 0.89, 0.91, and 0.94. 0.89, 0.92, respectively, with similar cutoff ranges 4.05 kPa, 4.7 kPa, and 5.3 kPa. These cut-offs were higher than those reported in Europe-USA in previous meta-analysis(26). We observed that both CAP and LSM cutoffs exhibited a statistically significant difference between each grade for normal and obese/overweight patients & normal and above normal WHR. This suggests that obesity/overweight and central obesity, as indicated by a raised WHR(28), may be an early indicator of liver steatosis and further fibrosis.

These strong positive correlation observed between the CAP and LSM(r=0.785, p<0.001) in our study corroborates findings from other research, highlighting the utility of these non-invasive metrics in assessing liver health(26,29). Specifically, the high R^2^ value of 0.616 suggests that CAP explains a substantial portion of the variability in LSM, underscoring its potential as a reliable indicator for comprehensive liver assessment. This robust correlation indicates that CAP, primarily used for steatosis quantification, provides valuable insights into the fibrotic changes reflected by LSM(30). The regression equation, LSM = 0.051 × CAP – 10, further quantifies this relationship, enabling a predictive model for liver stiffness based on steatosis measurements. This predictive capacity is particularly significant given that hepatic steatosis, while not directly causing fibrosis, often precedes steatohepatitis, which is strongly associated with progressive fibrotic changes(31). This interdependence emphasizes the value of concurrent assessment of both parameters for a more holistic evaluation of liver disease progression and risk stratification. This finding supports the integration of FibroScan into routine non-invasive screening protocols for MASLD, offering an early diagnostic opportunity for clinicians.

Of the risk factors for MASLD, in our study, there were a very small percentage of patients with history of diabetes (6.3%) and untreated dyslipidemia (5.5%). The majority of the participants with either obese/ overweight with a higher BMI (88.2%) and WHR(82.8%) making truncal adiposity, the most relevant predisposing factor to MASLD. The crucial highlight of this finding is that, while diagnosed metabolic conditions might be low at baseline, the underlying high rates of adiposity signify a substantial risk for MASLD development and progression. This aligns with global trends where the prevalence of MASLD is significantly increasing due to rising rates of obesity and metabolic syndrome(32,33). Danielsson et al. showed that high values of WHR are risk factors for severe liver disease, and both waist and hip circumference are independently associated with liver disease risk(34). Similarly, Aberg et al. found that the waist-hip ratio is superior to BMI in predicting liver-related outcomes(35). Fofiu et al. also linked visceral adiposity indices such as visceral adiposity index (VAI) and lipid accumulation product (LAP) to the prediction of hepatic steatosis and fibrosis in MASLD patients(36). Hao et al. additionally demonstrated the association of the WHR with MAFLD incidence(37). Our results reinforce the notion that, along with BMI, WHR should be considered in MASLD risk stratification and management strategies, especially for identifying individuals at higher risk of developing liver fibrosis. Duseja et al. recommend that MASLD case-finding approaches be directed towards individuals presenting with cardiometabolic risk factors, abnormal liver enzymes, or signs of hepatic steatosis on imaging, particularly when associated with type 2 diabetes or obesity alongside other metabolic risk factor(s)(38). Our findings stress the importance of early detection and intervention in this high-risk population, even before the manifestation of overt metabolic syndrome components.

## Strength & Limitations

A key strength of this study is calculated the sample size with good power. The study uses advanced VCTE with SmartExam underscores its commitment to modern and precise diagnostic methodologies. It significantly contributes to public health by aligning with global and national health priorities like UN SDG and 12. While the present study offers valuable insights into the relationship between CAP and LSM, the study cohort was drawn from a single center, the generalizability of its results to larger populations may be restricted, necessitating validation through multi-center studies.

## Conclusion

In conclusion, our study confirms the excellent performance of CAP in diagnosing and stratifying hepatic steatosis (for overall= 245.5 dB/m for ≥S1, 259.5 dB/m for ≥S2, and 275.5 dB/m for S3) and the promising role of LSM in assessing fibrosis (≥F1, ≥F2, and F3 were 3.95 kPa, 4.45 kPa, and 5.05kPa respectively) in the MASLD cohort. The strong positive correlation between the CAP and LSM highlights that the same diagnostic values can be used for LSM to measure liver stiffness. Furthermore, the findings emphasize that a diagnostic approach involving multiple diagnostic methods is essential even after automatic probe selection, particularly in patients with varying anthropometric parameters, where combining CAP and LSM with other NITs could probably enhance diagnostic precision for steatosis and fibrosis. This can be taken into consideration and implemented in the operational guidelines for MASLD under NPNCDs by the policy makers to identify and stratify the initial fibrotic stages.

## Data Availability

The participants' data that has been used is confidential. Data cannot be shared publicly because of patient confidentiality.

## Acknowledgements

The authors acknowledge the Dean, Medical Superintendent and Head of the department for their support throughout this research. Our heartfelt thanks go to all the dedicated technicians involved in this study for their meticulous work. Additionally, we extend my gratitude to the authors of the research works referenced in this study. Finally, and most importantly, we are profoundly grateful to all the participants in this study, whose willingness to contribute made this research possible.

## Supporting information

**Table S1:** Comparison of anthropometric parameters of Study participants in different grades (n=128)

| Biochemical parameters | S0<br>N=32 | S1<br>N=32 | S2<br>N=32 | S3<br>N=32 | P-value |
| --- | --- | --- | --- | --- | --- |
| BMI (kg/m<sup>2</sup>) | 24.38(3.58) | 27.87(3.83) | 29.81(4.09) | 29.90(4.51) | <0.001 |
| WHR | 0.87(0.05) | 0.91(0.06) | 0.97(0.05) | 0.98(0.03) | <0.001 |

S0, Steatosis 0; S1, steatosis 1; S2, steatosis 2; S3, steatosis 3; BMI, body mass index; WHR, waist-hip ratio

## Notes

### Competing Interest Statement

The authors have declared no competing interest.

### Funding Statement

The author(s) received no specific funding for this work.

### Author Declarations

Ethical approval was received from the institutional ethics committee (IECKMCMLR06/2024/346). Written informed consent was obtained from all participants prior to their inclusion in the study.

## References

1. Noncommunicable diseases [Internet]. [cited 2025 July 22]. Available from: https://www.who.int/health-topics/noncommunicable-diseases

2. Noncommunicable diseases India 2018 country profile [Internet]. [cited 2025 Aug 20]. Available from: https://www.who.int/publications/m/item/noncommunicable-diseases-ind-country-profile-2018

3. Asrih M, Jornayvaz FR. Metabolic syndrome and nonalcoholic fatty liver disease: Is insulin resistance the link? Mol Cell Endocrinol. 2015 Dec 15;418:55–65.

4. European Association for the Study of the Liver, European Association for the Study of Diabetes, European Association for the Study of Obesity. EASL–EASD–EASO Clinical Practice Guidelines on the management of metabolic dysfunction-associated steatotic liver disease (MASLD): Executive Summary. Diabetologia. 2024;67(11):2375–92.

5. Byrne CD, Targher G. NAFLD: A multisystem disease. J Hepatol. 2015 Apr 1;62(1, Supplement):S47–64.

6. Mantovani A, Csermely A, Petracca G, Beatrice G, Corey KE, Simon TG, et al. Non-alcoholic fatty liver disease and risk of fatal and non-fatal cardiovascular events: an updated systematic review and meta-analysis. Lancet Gastroenterol Hepatol. 2021 Nov 1;6(11):903–13.

7. Stefan N, Yki-Järvinen H, Neuschwander-Tetri BA. Metabolic dysfunction-associated steatotic liver disease: heterogeneous pathomechanisms and effectiveness of metabolism-based treatment. Lancet Diabetes Endocrinol. 2025 Feb 1;13(2):134–48.

8. Operational Guidelines for NALFD.cdr [Internet]. [cited 2025 Aug 28]. Available from: https://www.mohfw.gov.in/sites/default/files/Operational%20Guidelines%20for%20NALFD%20Version%2020.pdf

9. Khalifa A, Rockey DC. The Value of Liver Biopsy and Histology in Liver Disease Diagnosis and Patient Care-a Pragmatic Prospective Clinical Practice Study. J Clin Gastroenterol. 2024 Oct 1;58(9):912–6.

10. Elizabeth E Williams RV. Role of Vibration-Controlled Transient Elastography in the Evaluation and Management of Metabolic Dysfunction-Associated Steatotic Liver Disease. Curr Hepatol Rep. 2024;23:355–63.

11. Vuppalanchi R, Loomba R. Noninvasive Tests to Phenotype Nonalcoholic Fatty Liver Disease: Sequence and Consequences of Arranging the Tools in the Tool Box. Hepatology. 2021 June;73(6):2095.

12. Cao Y tian, Xiang L lan, Qi F, Zhang Y juan, Chen Y, Zhou X qiao. Accuracy of controlled attenuation parameter (CAP) and liver stiffness measurement (LSM) for assessing steatosis and fibrosis in non-alcoholic fatty liver disease: A systematic review and meta-analysis. eClinicalMedicine [Internet]. 2022 Sept 1 [cited 2025 May 26];51. Available from: https://www.thelancet.com/journals/eclinm/article/PIIS2589-5370(22)00277-2/fulltext

13. SDG Target 3.4 | Noncommunicable diseases and mental health: By 2030, reduce by one third premature mortality from non-communicable diseases through prevention and treatment and promote mental health and well-being [Internet]. [cited 2025 July 21]. Available from: https://www.who.int/data/gho/data/themes/topics/indicator-groups/indicator-group-details/GHO/sdg-target-3.4-noncommunicable-diseases-and-mental-health

14. Duseja A, Singh SP, D. A, Madan K, Rao PN, Shukla A, et al. Indian National Association for Study of the Liver (INASL) Guidance Paper on Nomenclature, Diagnosis and Treatment of Nonalcoholic Fatty Liver Disease (NAFLD). J Clin Exp Hepatol. 2023;13(2):273–302.

15. What Is A Standard Drink? | National Institute on Alcohol Abuse and Alcoholism (NIAAA) [Internet]. [cited 2024 May 9]. Available from: https://www.niaaa.nih.gov/alcohols-effects-health/overview-alcohol-consumption/what-standard-drink

16. Sanyal AJ, Brunt EM, Kleiner DE, Kowdley K, Chalasani N, Lavine J, et al. ENDPOINTS AND CLINICAL TRIAL DESIGN FOR NONALCOHOLIC STEATOHEPATITIS. Hepatol Baltim Md. 2011 July;54(1):344–53.

17. Misra A, Nigam P, Hills AP, Chadha DS, Sharma V, Deepak KK, et al. Consensus physical activity guidelines for Asian Indians. Diabetes Technol Ther. 2012 Jan;14(1):83–98.

18. Misra A, Misra R, Wijesuriya M, Banerjee D. The metabolic syndrome in South Asians: continuing escalation & possible solutions. Indian J Med Res. 2007 Mar;125(3):345–54.

19. Waist circumference and waist-hip ratio: report of a WHO expert consultation [Internet]. 2025 [cited 2025 June 8]. Available from: https://www.who.int/publications/i/item/9789241501491

20. Ethnic-Specific Criteria for Classification of Body Mass Index: A Perspective for Asian Indians and American Diabetes Association Position Statement | Diabetes Technology & Therapeutics [Internet]. [cited 2025 July 26]. Available from: https://www.liebertpub.com/doi/10.1089/dia.2015.0007

21. Hemant Kulkarni, Amit Amin, Manju Mamtani, Manik Amin. Diagnostic Test - OpenEpi module for performance evaluation of a diagnostic test [Internet]. [cited 2025 July 7]. Available from: https://www.openepi.com/DiagnosticTest/DiagnosticTest.htm

22. Kumar M, Rastogi A, Singh T, Behari C, Gupta E, Garg H, et al. Controlled attenuation parameter for non-invasive assessment of hepatic steatosis: Does etiology affect performance? J Gastroenterol Hepatol. 2013;28(7):1194–201.

23. Karlas T, Petroff D, Sasso M, Fan JG, Mi YQ, de Lédinghen V, et al. Individual patient data meta-analysis of controlled attenuation parameter (CAP) technology for assessing steatosis. J Hepatol. 2017 May 1;66(5):1022–30.

24. Body mass index–based controlled attenuation parameter cut-offs for assessment of hepatic steatosis in non-alcoholic fatty liver disease | Indian Journal of Gastroenterology [Internet]. 2025 [cited 2025 June 27]. Available from: https://link.springer.com/article/10.1007/s12664-019-00991-2

25. Fujiwara Y, Kuroda H, Abe T, Nagasawa T, Nakaya I, Ito A, et al. Impact of shear wave elastography and attenuation imaging for predicting life-threatening event in patients with metabolic dysfunction-associated steatotic liver disease [Internet]. Research Square; 2024 [cited 2025 July 26]. Available from: https://www.researchsquare.com/article/rs-5162272/v1

26. Cao Y tian, Xiang L lan, Qi F, Zhang Y juan, Chen Y, Zhou X qiao. Accuracy of controlled attenuation parameter (CAP) and liver stiffness measurement (LSM) for assessing steatosis and fibrosis in non-alcoholic fatty liver disease: A systematic review and meta-analysis. eClinicalMedicine [Internet]. 2022 Sept 1 [cited 2025 May 26];51. Available from: https://www.thelancet.com/journals/eclinm/article/PIIS2589-5370(22)00277-2/fulltext

27. Chouik Y, Aubin A, Maynard-Muet M, Segrestin B, Milot L, Hervieu V, et al. The grade of obesity affects the noninvasive diagnosis of advanced fibrosis in individuals with MASLD. Obesity. 2024;32(6):1114–24.

28. Baioumi AYAA. Chapter 3 - Comparing Measures of Obesity: Waist Circumference, Waist-Hip, and Waist-Height Ratios. In: Watson RR, editor. Nutrition in the Prevention and Treatment of Abdominal Obesity (Second Edition) [Internet]. Academic Press; 2019 [cited 2025 Sept 3]. p. 29–40. Available from: https://www.sciencedirect.com/science/article/pii/B9780128160930000033

29. Fujiwara Y, Kuroda H, Abe T, Nagasawa T, Nakaya I, Ito A, et al. Impact of shear wave elastography and attenuation imaging for predicting life-threatening event in patients with metabolic dysfunction-associated steatotic liver disease [Internet]. Research Square; 2024 [cited 2025 Aug 14]. Available from: https://www.researchsquare.com/article/rs-5162272/v1

30. Ding Y, Wang Z, Niu H, Deng Q, Wang Y, Xia S. FIB-4 is closer to FibroScan screen results to detecting advanced liver fibrosis and maybe facilitates NAFLD warning. Medicine (Baltimore). 2023 Aug 25;102(34):e34957.

31. Soman DrN, Kharat DrA, Jesson DrJ. Evaluation of Liver Stiffness using Magnetic Resonance Elastography in Hepatic Steatosis. Sch J Appl Med Sci. 2021 Mar 28;9(3):462–7.

32. Miao L, Targher G, Byrne CD, Cao YY, Zheng MH. Current status and future trends of the global burden of MASLD. Trends Endocrinol Metab. 2024 Aug 1;35(8):697–707.

33. Kan C, Zhang K, Wang Y, Zhang X, Liu C, Ma Y, et al. Global burden and future trends of metabolic dysfunction-associated Steatotic liver disease: 1990-2021 to 2045. Ann Hepatol. 2025 July 1;30(2):101898.

34. Waist and hip circumference are independently associated with the risk of liver disease in population-based studies - Danielsson - 2021 - Liver International - Wiley Online Library [Internet]. [cited 2025 July 26]. Available from: https://onlinelibrary.wiley.com/doi/10.1111/liv.15053

35. Åberg F, Färkkilä M, Salomaa V, Jula A, Männistö S, Perola M, et al. Waist-hip ratio is superior to BMI in predicting liver-related outcomes and synergizes with harmful alcohol use. Commun Med. 2023 Sept 6;3(1):119.

36. Bende R, Heredea D, Raţiu I, Sporea I, Dănilă M, Șirli R, et al. Association Between Visceral Adiposity and the Prediction of Hepatic Steatosis and Fibrosis in Patients with Metabolic Dysfunction-Associated Steatotic Liver Disease (MASLD). J Clin Med. 2025 Jan;14(10):3405.

37. Waistline to thigh circumference ratio as a predictor of MAFLD: a health care worker study with 2-year follow-up | BMC Gastroenterology | Full Text [Internet]. [cited 2025 July 26]. Available from: https://bmcgastroenterol.biomedcentral.com/articles/10.1186/s12876-024-03229-4

38. Duseja A, Singh SP, D. A, Madan K, Rao PN, Shukla A, et al. Indian National Association for Study of the Liver (INASL) Guidance Paper on Nomenclature, Diagnosis and Treatment of Nonalcoholic Fatty Liver Disease (NAFLD). J Clin Exp Hepatol. 2023;13(2):273–302.

